# The Effect of the COVID-19 Pandemic on People with Parkinson’s Disease

**DOI:** 10.1101/2020.07.14.20153023

**Authors:** Ethan G. Brown, Lana M. Chahine, Samuel M. Goldman, Monica Korell, Emerald Mann, Daniel R. Kinel, Vanessa Arnedo, Kenneth L. Marek, Caroline M. Tanner

## Abstract

**Objective:** To rapidly identify areas of need and improve care in people with Parkinson’s disease (PwPD) affected by the COVID-19 pandemic, we deployed a survey focusing on the presentation and complications of COVID-19 infection and the effect of the COVID-19 pandemic among those not infected.

**Methods:** Individuals with and without PD participating in the online study Fox Insight (FI) were invited to complete an online survey between April 23-May 23, 2020. Among people reporting COVID-19 diagnosis, we compared the frequency of symptoms and poor outcomes in people with and without PD. Among people not reporting COVID-19, we assessed the effects of the pandemic on access to medical care and other services, and in PwPD, its effects on PD symptoms.

**Results:** Among 5,429 PwPD and 1,452 without PD, 77 reported a COVID-19 diagnosis (51 PwPD, 26 without PD). Complications were more frequent in people with longer PD duration. PwPD and COVID-19 experienced new or worsening motor (63%) and nonmotor (75%) PD symptoms. PwPD not diagnosed with COVID-19 reported disrupted medical care (64%), exercise (21%), and social activities (57%), and worsened PD motor (43%) and non-motor (52%) symptoms. Disruptions were more common for PwPD living alone, and for those with lower income and non-white race.

**Conclusions:** The COVID-19 pandemic is associated with wide-ranging effects on people with PD, and certain groups may be at particular risk. FI provides a rapid, patient-centered means to assess these effects and identify needs that can be used to improve the health of PwPD.

## Introduction

SARS-CoV-2 infection and the societal changes associated with the COVID-19 pandemic may have particularly severe consequences for people with chronic neurologic diseases, such as Parkinson’s disease (PD). Many people with PD (PwPD) are concerned about COVID-19 risk,^1^ and indeed early reports^2-4^ describe worsening of parkinsonian symptoms during infection and poor outcomes in some cases. Among those not infected, social distancing guidelines may lead to social isolation, a known risk factor for poor outcomes,^5^ and reduced access to care,^6^ all of which are additional concerns of PwPD.^1^ Rapid changes in healthcare delivery to PwPD have been developed, including recommendations on implementing telemedicine^7^ and managing advanced therapies remotely,^8, 9^ but these resources are not universally available.^10^ The persistent spread of the pandemic and the associated global disruption highlight the urgent need for increased understanding of its effect and mitigating strategies in PwPD.

We surveyed participants in Fox Insight (FI), an online study that involves thousands of people with and without PD,^11^ between April and May, 2020. Our goals were to (1) understand the symptoms and outcomes of SARS CoV-2 infection in people with and without PD, (2) determine its effects on PD symptoms; and (3) understand the effects of the pandemic and associated public health measures. We were particularly interested in how demographic factors, including age, sex, and socioeconomic factors, related to disruptions from the pandemic.

## Methods

### Description of Fox Insight

Fox Insight (FI) is a fully remote study platform.^12^ Participants ≥18 years old with and without PD complete surveys at regular intervals and are periodically invited to complete additional surveys targeting particular topics.^11^

### Survey design

This was a cross-sectional study. The COVID-19 survey was designed through an iterative process involving multidisciplinary providers and clinical researchers with active feedback on survey content and format from PwPD.

### Study population and survey deployment

All FI participants were invited by email on April 23, 2020, with follow-up reminders on May 1^st^ and May 11^th^ to participate in the COVID-19 survey. Because spread of the pandemic and related attitudes, guidelines, and practices have changed rapidly, we limited analyses to surveys completed between April 23^rd^ and May 23^rd^, 2020. We excluded respondents with missing diagnostic or demographic data in FI and those who reported a diagnosis of PD before age 25.

### Assessments

Participant information comes both from FI longitudinal study assessments and direct questions of the COVID-19 survey. From FI, we obtained demographic information that participants provide annually. For covariate analysis, older age was defined as more than 65 years at completion of the survey. Respondents of “non-white race” include those who did not identify as White and identified as another race (either African American, Asian, American Indian or Alaska Native, Native Hawaiian or Pacific Islander, according to the choices in FI). Participants with a household income below $50,000, representing roughly the lowest tertile of respondents, were classified as having lower income. Several PD-related genes may be pertinent to COVID-19 infection: LRRK2 mutations may play a role in immunomodulation,^13^ while APOE ε4 homozygosity has been associated with increased COVID-19 risk^14^ and, along with GBA, may increase susceptibility to cognitive impairment in PwPD.^15^ Genotype was therefore included for the subset of PwPD in FI who had those data available.^11^ Information about PD diagnosis was obtained both from FI and from our current survey; respondents with conflicting answers were excluded.

All subsequent assessments of COVID-19 and the effect of the pandemic were obtained through the deployed survey. We considered people to have COVID-19 if they reported that a definite or probable COVID-19 diagnosis had been made by a medical professional. People reporting a history of diabetes, HIV or AIDS, recent chemotherapy, current treatment with oral steroids or other immune suppressants were classified as immunocompromised. To understand how the pandemic is affecting both infected and non-infected people with and without PD, the survey was divided into three sections: (1) COVID-19 symptoms, diagnosis, testing, risk factors, and treatment; (2) change in PD-related symptoms; and (3) effect of shelter-in-place orders and social distancing practices on healthcare access, social interactions, and other essential activities (the full survey is available online).^16^

### Definition of outcomes

Outcomes assessed in people with COVID-19 included development of pneumonia, need for supplemental oxygen, hospitalization, intensive care unit (ICU) admission or ventilator support. For people without COVID-19, we assessed pandemic-associated disruption of medical care, daily essential activities, and exercise or social activities. We evaluated changes in individual PD-related motor and non-motor symptoms by domain: motor problems (imbalance, falling, tremor, slow movement, stiffness, dysphagia, difficulty eating, increased OFF time, dyskinesia), cognitive problems (confusion, hallucinations, difficulty thinking, memory problems), sleep problems (acting out dreams, difficulty sleeping, excessive sleepiness, fatigue), mood symptoms (anxiety, depression, apathy), and autonomic problems (urinary problems, orthostasis, constipation).

### Statistical analysis

Survey results and demographic information were summarized using descriptive statistics. We evaluated differences in means of continuous variables using Student’s t-test for normally distributed data and Wilcoxon rank sum for non-normally distributed. We used the chi-square test for comparison of categorical variables. For our main results, we constructed logistic regression models adjusting for age and sex. For COVID-19 related analyses, we performed sensitivity analyses restricting the cohort to only those reporting a definite COVID-19 diagnosis or a positive COVID-19 test. Participants who reported COVID-19 infection were excluded from analyses about the effects of social distancing to reduce confounding. All analyses were performed using R version 3.5.2 (12-20-2018). Statistical significance was defined as p<0.05.

### Standard Protocol Approvals, Registrations, and Patient Consents

The FI study and the survey for this COVID-19 study were approved by the New England IRB and informed consent was obtained online from all participants.

### Data Availability Policy

All survey questions and data are available for download online.^16^

## Results

As of May 23, 2020, 7,209 people with and without PD responded, representing more than 50% of the 14,166 Fox Insight participants active in the preceding 12 months. Three-hundred and twenty-eight people were excluded for irreconcilable or missing data, leaving 5,429 people with PD and 1,452 people without. Responses were from all continents, but about 80% came from the United States (**Figure 1**). Respondents with PD were older, more often female, had a higher prevalence of heart disease, a lower prevalence of immunocompromising conditions and lung disease, and had lower household income than people without PD (**Table 1**).

**Table 1:** Characteristics of respondents. Numbers in parentheses represent range for continuous variables and percentage of specific cohort for categorical variables. Immunocompromised defined as having a history of diabetes, HIV or AIDS, chemotherapy within the past year, taking steroid medications by mouth, or taking other immune suppressing medication. Note: genotype was not available for people without PD.

|  | COVID-19 Infection |  |  | No COVID-19 Infection |  |  |
| --- | --- | --- | --- | --- | --- | --- |
|  | PD<br>N = 51 | Not PD<br>N = 26 | p -<br>value* | PD<br>N = 5378 | Not PD<br>N = 1426 | p -<br>value |
| Age, mean in years (range) | 65 (40 - 89) | 57 (30 - 73) | **0.004** | 68 (33 - 95) | 61(19 - 94) | **<0.001** |
| Female, N (%) | 27 (53) | 24 (92) | **0.001** | 2598 (48) | 1115 (78) | **<0.001** |
| PD Duration, N (%) |  |  |  |  |  |  |
| 0 – 3y | 21 (41) | -- |  | 1628 (30) | -- |  |
| 3 – 6y | 12 (24) | -- |  | 1649 (31) | -- |  |
| 6 – 9y | 9 (18) | -- |  | 981 (18) | -- |  |
| > 9y | 9 (18) | -- |  | 1114 (21) | -- |  |
| Comorbidities, N(%) |  |  |  |  |  |  |
| Immunocompromised | 11 (22) | 8 (31) | 0.5 | 637 (12) | 198 (14) | **0.041** |
| Current Smoker | 3 (5.9) | 0 (0) | 0.5 | 84 (1.6) | 32 (2.2) | 0.10 |
| Former Smoker | 20 (39) | 7 (27) | 0.4 | 1408 (26) | 368 (26) | 0.8 |
| Heart Disease | 10 (20) | 1 (3.8) | 0.087 | 440 (8.2) | 90 (6.3) | **0.022** |
| Hypertension | 13 (25) | 8 (31) | 0.8 | 1649 (31) | 439 (31) | >0.9 |
| Lung disease | 7 (14) | 11 (42) | **0.012** | 436 (8.1) | 198 (14) | **<0.001** |
| White, N (%) | 51 (100) | 26 (100) |  | 5247 (98) | 1396 (98) | 0.5 |
| Latinx, N (%) | 4 (8.0) | 0 (0) | 0.3 | 168 (3.2) | 42 (3.0) | 0.7 |
| Income < \$50,000, N (%) | 13 (28) | 6 (25) | >0.9 | 1201 (26) | 274 (22) | **0.002** |
| Lives Alone, N (%) | 8 (19) | 4 (17) | >0.9 | 630 (13) | 256 (19) | **<0.001** |
| Genotype, N (% with genotype data) |  |  |  |  |  |  |
| APOE ε4 allele (n=599) |  |  |  |  |  |  |
| None | 3 (100) | -- |  | 450 (76) | -- |  |
| One | 0 | -- |  | 135 (23) | -- |  |
| Two | 0 | -- |  | 7 (1.2) | -- |  |
| GBA Mutation (n=609) | 0 | -- |  | 39 (6.5) | -- |  |
| LRRK2 G2019S (n=640) | 0 | -- |  | 28 (4.4) | -- |  |

\*p-values represent comparisons between people with and without PD.

**Figure 1:**
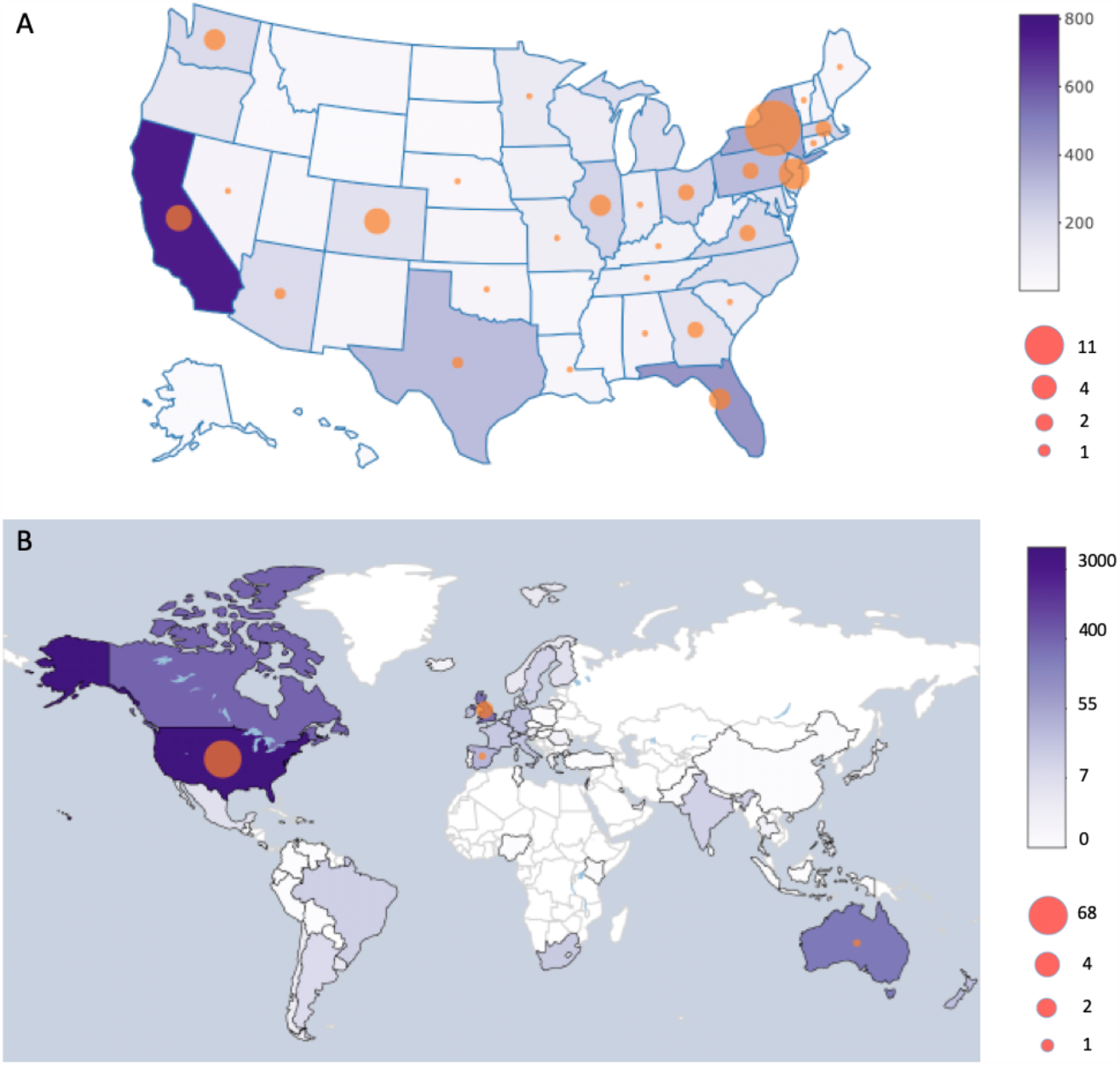
Distribution of survey responses and COVID-19 cases among respondents (A) within the US and (B) across the globe. The color of the state or country represents the number of survey responses and the size of the circles represents the number of COVID-19 diagnoses reported from each region.

COVID-19 diagnoses were reported by 51 PwPD (22 definite, 29 probable) and 26 people without PD (7 definite, 19 probable). Positive tests were reported in 17 PwPD and 6 without PD. One PwPD reported a positive COVID-19 test but was asymptomatic and was not included in analyses of COVID-19-associated disease features. Among PwPD, those with COVID-19 were more likely to smoke (5.9% vs 1.6%, p = 0.048) and have a history of heart disease (20% vs 8.2%, p = 0.008). Compared to people without PD who had COVID-19, PwPD who had COVID-19 were more likely to be older, male, and less likely to have lung disease (**Table 1**).

Behavioral and environmental risk factors for COVID-19 were more common in people without PD than PwPD: 100% of people without PD with COVID-19 reported a professional or recreational activity that potentially put them at risk, whereas only 75% of PwPD with COVID-19 could identify any such activity (p = 0.012). PwPD and COVID-19 were more likely to have a household or other personal contact that was diagnosed with, suspected to have, or had symptoms consistent with COVID-19 compared to PwPD without COVID-19 (59% vs 7.7%, p < 0.001). The types of COVID-19 symptoms were generally similar in PwPD and those without except for more frequent chills and pulmonary symptoms in people without PD (**Table 2**). Among people with COVID-19, the new onset of hyposmia was reported by 3.9% of PwPD and 7.7% of people without PD, and worsening of existing hyposmia was reported by 26% of PwPD and 19.2% of those without PD. Among people without COVID-19, new onset of hyposmia was reported by 0.4% of PwPD and 0.2% of people without PD, and worsening of existing hyposmia was reported by 3.1% of PwPD and 3.1% of people without PD. Among people with COVID-19, there were slight differences in several reported outcomes between those with and without PD, but none were statistically significant (**Table 2**). Longer PD-duration was associated with a higher risk of pneumonia, the need for supplemental oxygen, or hospitalization (44% among PwPD for greater than 9 years vs 14% among PwPD with equal to or less than 9 years, aOR 5.44, 95% CI 1.04 - 30.5, p= 0.043).

**Table 2:** Symptoms and outcomes of COVID-19 in people with and without PD. P-value represents results of chi-square test. Upper respiratory symptoms include congestion and sore throat. Lower respiratory symptoms include chest tightness, chest pain, and shortness of breath. GI symptoms include nausea, vomiting, diarrhea, and stomach pain. O2: oxygen. ICU: intensive care unit.

| **COVID-19 Symptoms and Outcomes in People with and without PD** |  |  |  |
| --- | --- | --- | --- |
| **Symptoms** |  |  |  |
|  | **PD, N = 51** | **Not PD, N = 26** | **p-value** |
| Fever | 32 (63%) | 20 (77%) | 0.3 |
| Chills | 28 (55%) | 21 (81%) | **0.048** |
| Cough | 36 (71%) | 25 (96%) | **0.020** |
| Upper Respiratory | 35 (69%) | 20 (77%) | 0.6 |
| Lower Respiratory | 29 (57%) | 24 (92%) | **0.004** |
| GI | 34 (67%) | 20 (77%) | 0.5 |
| Muscle/Joint Pain | 39 (76%) | 24 (92%) | 0.2 |
| Sleepiness | 44 (86%) | 26 (100%) | 0.12 |
| Lightheadedness | 34 (67%) | 23 (88%) | 0.074 |
| **Outcomes** |  |  |  |
| Pneumonia | 4 (7.8%) | 3 (12%) | >0.9 |
| Supplemental O2 | 6 (12%) | 2 (7.7%) | 0.9 |
| Hospitalized | 5 (9.8%) | 2 (7.7%) | >0.9 |
| ICU | 2 (40%) | 1 (50%) | >0.9 |
| Ventilator | 1 (20%) | 0 (0%) | >0.9 |

During COVID-19 infection, PwPD reported worsening of many PD-related symptoms **(Figure 2)**. New motor symptoms were reported by 18%, and 55% reported worsening of at least one existing motor symptom. New and worsening non-motor symptoms were reported for all domains: mood (20% new, 51% worsening), cognition (7.8% new, 41% worsening), sleep (12% new, 59% worsening), and autonomic (7.8% new, 29% worsening).

**Figure 2:**
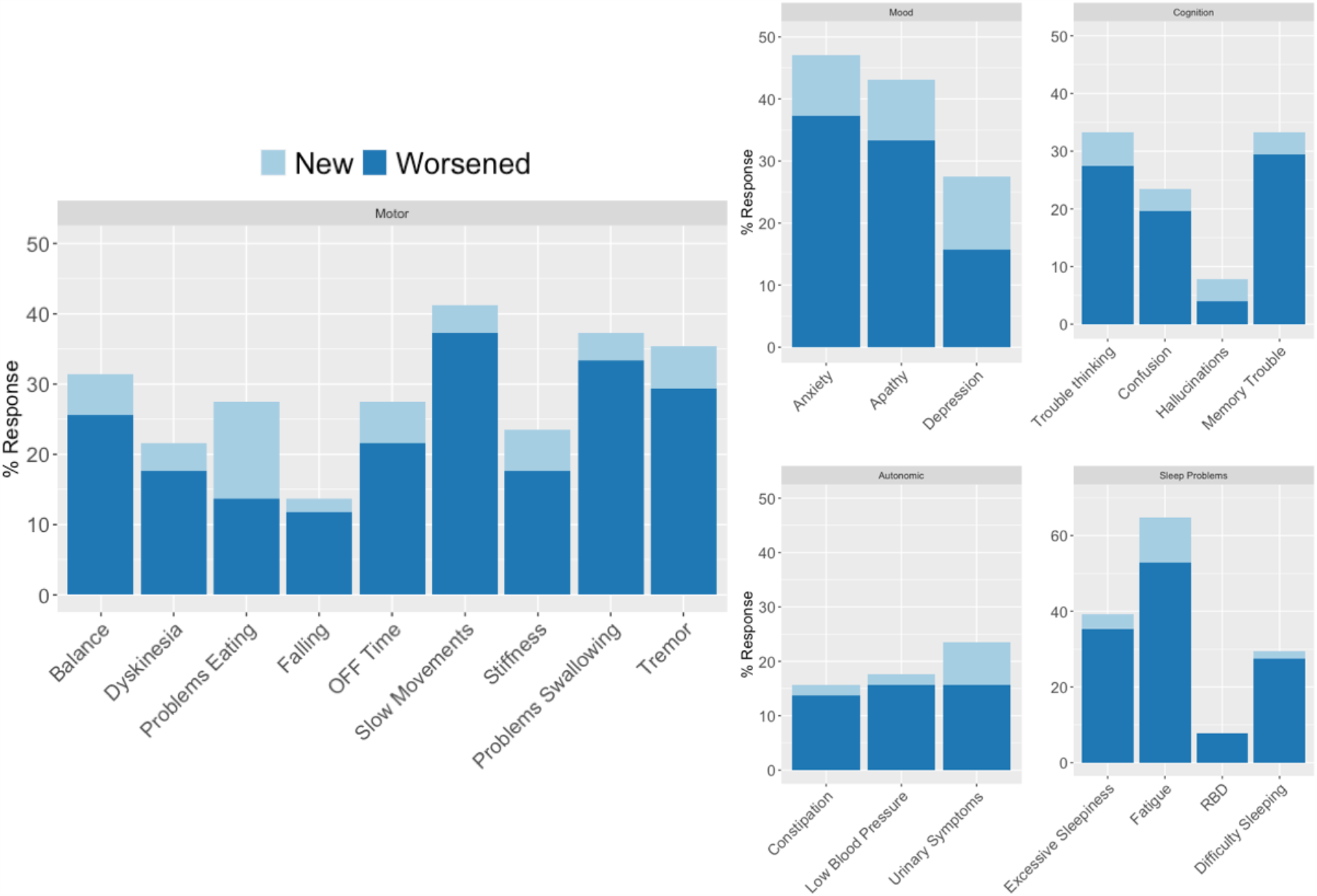
Changes in PD symptoms in PwPD and COVID-19 (n = 51). Changes in motor and non-motor PD-related symptoms among respondents with PD and COVID-19 (n = 51).

Among those without COVID-19, healthcare was altered due to the pandemic in 62% of all respondents, including cancelled appointments, reduced in-home care or difficulty obtaining medications. Only cancellations in rehabilitation services were more common in PwPD (17% PwPD vs 5.5% not PD, p < 0.001). Among PwPD, disruption in medical care was more likely in those with longer PD duration (41% PD duration > 9 years vs. 32% for PD duration 0-3 years, adjusted odds ratio (aOR) = 1.47, 95% CI 1.26 – 1.73, p < 0.001) (**Table 3**). Race and lower income were independently associated with difficulty obtaining PD medications (non-White race 13% vs 7.3%, aOR 1.98 95% CI 1.05 - 3.45, p = 0.023, and lower income 10% vs 7.2%, aOR 1.36, 95% CI 1.07 - 1.72, p = 0.01); arguably more people of Latinx ethnicity reported difficulty obtaining medications (14% vs 7.5%, 1.61, 95% CI 0.93 – 2.63, p = 0.07). Telemedicine appointments were reported by 39% of PwPD, but those with lower household income were less likely to attend healthcare appointments through telemedicine (40% vs 35%, aOR 0.79, 95% CI 0.69, 0.90, p = <0.001).

**Table 3:** Interruptions in PD-related medical care stratified by disease duration among people with PD without COVID-19 infection.

| **Disruptions of Medical Care in People with PD Without COVID-19 Related to the COVID-19 Pandemic** |  |  |  |  |  |
| --- | --- | --- | --- | --- | --- |
| **PD Duration (y)** | **0 – 3**<br>**N = 1628** | **3 – 6**<br>**N = 1649** | **6 – 9**<br>**N = 981** | **> 9**<br>**N = 1114** | **p-value<sup>2</sup>** |
| Cancelled or postponed rehab therapy | 396 (24%) | 426 (26%) | 265 (27%) | 331 (30%) | **0.016** |
| Have lost or reduced in-home care services | 35 (2.1%) | 47 (2.9%) | 35 (3.6%) | 69 (6.2%) | **<0.001** |
| Had to cancel healthcare appts | 771 (47%) | 813 (49%) | 511 (52%) | 593 (53%) | **0.011** |
| Cancelled or postponed mental health care | 78 (4.8%) | 68 (4.1%) | 28 (2.9%) | 48 (4.3%) | 0.12 |
| Problems obtaining meds for PD | 118 (7.2%) | 114 (6.9%) | 83 (8.5%) | 96 (8.6%) | 0.3 |
| Cancelled or postponed Botox Treatment | 21 (1.3%) | 47 (2.9%) | 25 (2.5%) | 45 (4.0%) | **<0.001** |
| Cancelled or postponed DBS Surgery | 7 (0.4%) | 10 (0.6%) | 16 (1.6%) | 14 (1.3%) | **0.004** |
| Cancelled or postponed DBS Battery Replacement | 0 (0%) | 1 (<0.1%) | 3 (0.3%) | 7 (0.6%) | **0.002** |
| Cancelled or postponed DBS programming | 3 (0.2%) | 14 (0.8%) | 22 (2.2%) | 54 (4.8%) | **<0.001** |

At least one essential daily activity was disrupted in 35% of PwPD. Essential daily activities were more often disrupted in PwPD who live alone compared to other PwPD, including getting food (12% vs 8.7%, aOR 1.50, 95% CI 1.14 – 1.94, p = 0.003) and getting regular homecare / housekeeping 15% vs 9.0%, aOR 1.50, 95% 1.16 - 1.91, p = 0.001). Other regular behaviors disrupted due to the COVID-19 pandemic included cancelled exercise (21% PwPD) and social activities (57%), although many people found alternative ways, such as online classes, to continue these activities (**Table 4**). PwPD with lower income were less likely to report alternative means of exercising (32% vs 44%, aOR 0.57, 0.50 – 0.66, p<0.001) or social activities (49% vs 58%, aOR 0.66, 0.58 – 0.76, p <0.001); older PwPD were less likely to use alternative ways to exercise (39% vs 44%, aOR 0.82 (95% CI 0.73, 0.93), p=0.002).

**Table 4:** Interruptions in activities during the COVID-19 pandemic among people with PD but not infected by COVID-19.

| Changes in activities among people with PD not infected by COVID |  |  |  |  |
| --- | --- | --- | --- | --- |
|  | Cancelled | Postponed | Conducted via alternative method(s) | Not applicable |
| Exercise | 1140<br>(21%) | 427<br>(7.9%) | 2187<br>(41%) | 1624<br>(30%) |
| Seeing Family | 1316<br>(24%) | 1175<br>(22%) | 2062<br>(38%) | 825<br>(15%) |
| Seeing Friends | 1572<br>(29%) | 1341<br>(25%) | 1925<br>(36%) | 540<br>(10%) |
| Support Group Attendance | 893 (17%) | 240<br>(4.5%) | 529<br>(9.8%) | 3716<br>(69%) |
| Volunteer Activities | 1062<br>(20%) | 459<br>(8.5%) | 423<br>(7.9%) | 3434<br>(64%) |
| Religious Activities | 1095<br>(20%) | 390<br>(7.3%) | 1266<br>(24%) | 2627<br>(49%) |
| Community Activities | 2156<br>(40%) | 940 (17%) | 620 (12%) | 1662<br>(31%) |

The pandemic was also disruptive to research participation among the cohort. Among the 16% of PwPD without COVID-19 who had been participating in research, 40% had to cancel and 35% had to postpone in-person research visits, while 25% were able to conduct research visits through alternative methods. Overall, 6.3% of PwPD felt that the COVID-19 pandemic had made them more likely to participate in research, while 11% felt that the pandemic had made them less likely to participate, and 83% reported no change in their likelihood to participate in research.

Among PwPD who did not have COVID-19, new PD symptoms emerged and existing symptoms worsened in all major domains (motor: 6.2% new, 41% worsened; mood: 6.5 % new, 30 % worsened; cognitive: 2.5 % new, 16 % worsened; sleep: 4.5% new, 32% worsened; autonomic: 2.6% new, 18% worsened (**Figure 3**). Respondents who experienced interruptions to PD-related medical care were also more likely to experience new or worsening symptoms in all domains (motor symptoms: 56% vs 36%, aOR 2.21, 95% CI 1.97 – 2.48, p<0.001; cognitive problems: 24% vs 14%, aOR 1.92, 95% CI 1.66-2.22, p<0.001; mood symptoms: 42% vs 30%, aOR 1.70, 95% CI 1.51-1.92, p <0.001; autonomic symptoms: 27% vs 17%, aOR 1.75, 95% CI 1.53-2.00, p <0.001; and sleep problems: 42% vs 31%, aOR 1.60, 95% CI 1.42-1.79 p <0.001). New onset motor symptoms in particular were more likely in those that had disruption of medical care (8.2% vs 5.1% aOR 1.63, 95% CI 1.31-2.04, p < 0.001). Respondents who experienced interruptions to exercise, social activities or were asked to self-isolate were also more likely to report worsening of PD symptoms (**Table 5**). Neither the number of APOE ε4 alleles nor PD-associated mutations in LRRK2 were associated with an increased risk of being diagnosed with COVID-19, and the number of APOE ε4 alleles and GBA mutations were not associated with new or worsening PD-related symptoms during the pandemic.

**Table 5:** Risk of new or worsening PD-related symptoms in various domains after cancelled or postpone exercise, cancelled or postponed social activities, or being asked to self-isolate / quarantine during the COVID-19 pandemic among people with PD and without COVID-19. PD-symptom domains include motor (problems with walking, balance, falling, tremor, slow movements, stiffness, swallowing, eating, increased off-time, or dyskinesia), cognition (problems with thinking, memory, confusion, hallucinations), mood (anxiety, depression, apathy), autonomic (constipation, urinary problems, low blood pressure), and sleep (insomnia, fatigue, excessive sleepiness, REM sleep behavior disorder). Odds ratios are shown with 95% confidence interval in parentheses. Odds ratios are adjusted for the other factors presented. *p<0.01, **p<0.001

| **Factors Associated with Worsening PD-related Symptoms during COVID-19 Pandemic among PwPD without COVID-19** |  |  |  |  |  |
| --- | --- | --- | --- | --- | --- |
|  | **Motor** | **Cognitive** | **Mood** | **Autonomic** | **Sleep** |
| Exercise Activities<br>Cancelled/<br>Postponed | 1.31**<br>(1.16, 1.49) | 1.28*<br>(1.10, 1.50) | 1.21*<br>(1.07, 1.38) | 1.23*<br>(1.06, 1.42) | 1.34**<br>(1.18, 1.52) |
| Social Activities<br>Cancelled/<br>Postponed | 1.69**<br>(1.47, 1.95) | 1.41**<br>(1.17, 1.72) | 1.63**<br>(1.40, 1.90) | 1.46**<br>(1.22, 1.76) | 1.31**<br>(1.13, 1.51) |
| Asked to self-isolate/<br>quarantine | 1.78**<br>(1.49, 2.13) | 1.90**<br>(1.55, 2.32) | 1.69**<br>(1.41, 2.02) | 1.81**<br>(1.48, 2.19) | 1.71**<br>(1.43, 2.04) |

**Figure 3:**
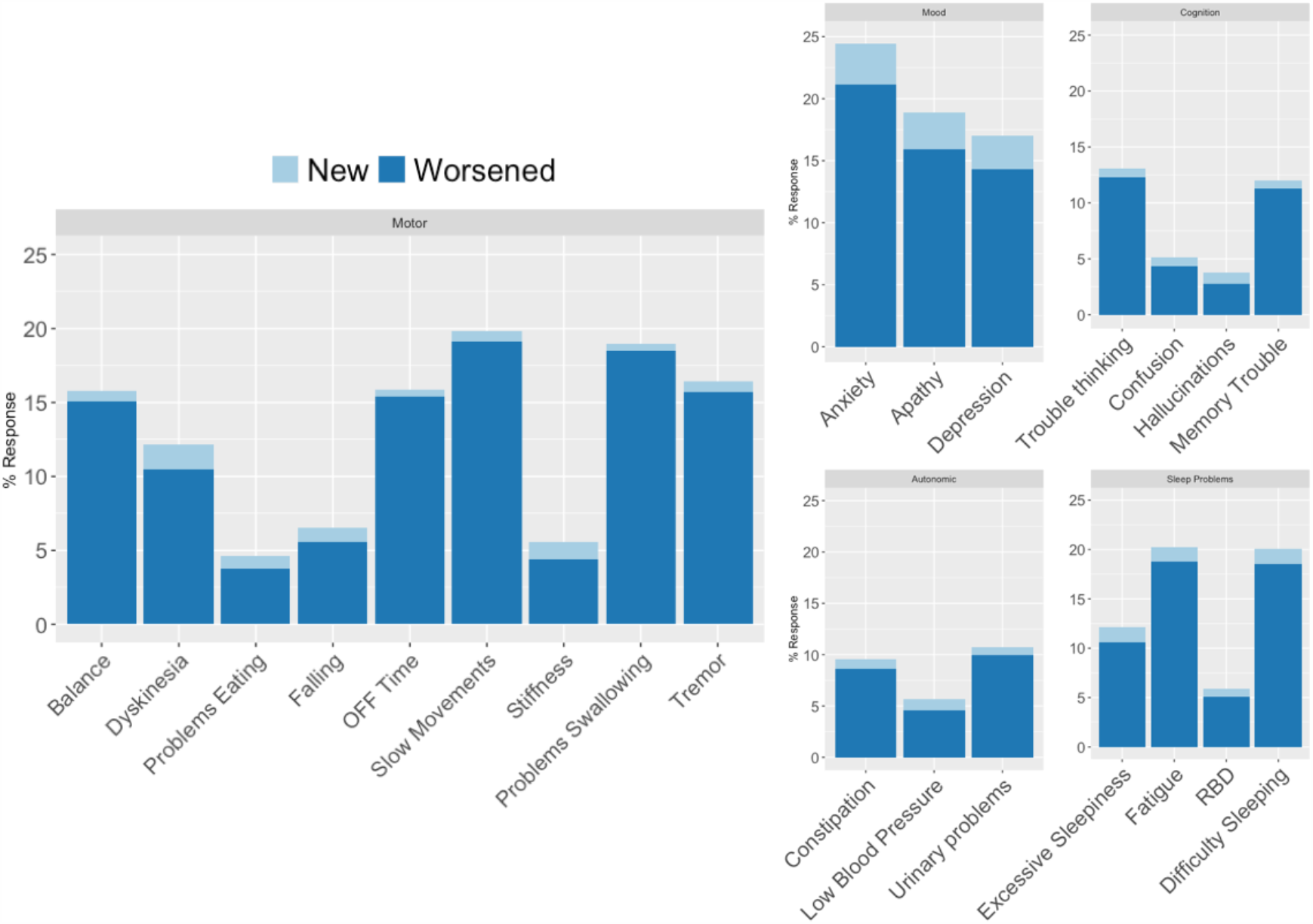
Changes in PD symptoms in PwPD without COVID-19 (n = 5,378). Changes in motor and non-motor PD-related symptoms among respondents with PD without COVID-19 (n = 5,378).

Women with COVID-19 more often reported new or worsening cognitive symptoms (67% women vs 25% men, aOR 6.00, 95% CI 1.82 – 22.3, p = 0.005). New or worsening symptoms were also more common in women with PD but without COVID-19: motor symptoms (49% women vs 38% men, aOR 1.63, 95% CI 1.46 - 1.81. p <0.001), mood symptoms (42% women vs 27% men, aOR 1.89, 95% CI 1.69 – 2.12, p<0.001), sleep problems (39% women vs 31% men, aOR 1.43, 95% CI 1.28 - 1.60, p <0.001), and autonomic problems (23% women vs 18% men, aOR 1.33, 95% CI 1.17 – 1.52, p < 0.001). In analyses adjusting for social factors more commonly experienced by women with PD (living alone, disrupted home care, interruptions in medical care), women with and without COVID-19 were still at greater risk of symptom worsening.

## Discussion

This large, participant reported study demonstrates the profound effect COVID-19 has had on individuals with PD. Thousands of people completed the survey in just a few weeks, illustrating the value of online research in providing rapid responses from an engaged community despite limits to in-person interactions.

Both motor and non-motor symptoms of PD worsened during SARS-CoV-2 infection, including stiffness, tremor, difficulty walking, mood symptoms, cognition, and fatigue. Others have similarly reported worsening of PD-related symptoms either as presenting signs of COVID-19^4^ or during the course of the infection.^2, 3^ Worsening of PD symptoms is commonly experienced in PwPD who have infectious illnesses,^17^ possibly due to systemic inflammation, altered dopaminergic signaling, or changes in medication absorption or pharmacokinetics.^18^ Worsening of symptoms due to a direct infection of the CNS by SARS-CoV-2 is less likely. Although COVID-19 has been associated with changes on neuroimaging^19, 20^ and SARS-CoV-2 RNA has been detected in cerebrospinal fluid,^21, 22^ a recent autopsy series of patients that died with COVID-19 — all of whom experienced altered mental status — did not find immunohistochemical evidence of encephalitis or viral invasion into brain tissue.^23^ Exacerbation of PD symptoms during COVID-19 may in part be due to the inflammatory reaction characteristic of the disease.^24^ The consistent reporting of symptom exacerbation in PwPD due to COVID-19 emphasizes the need to consider COVID-19 as a possible explanation for suddenly worsening PD-related symptoms.

Hyposmia has been well described in people with COVID-19.^25^ Although objective testing was not performed, respondents with and without PD who reported COVID-19 also reported new onset hyposmia. Interestingly, even PwPD who already had hyposmia reported worsening of this symptom. A minority of people without COVID-19 also experienced worsening of sense of smell, possibly representing undiagnosed SARS-CoV-2 infection.^26^

A greater proportion of women than men with PD reported COVID-19. Other case series of PwPD and COVID-19 have not shown an excess of women,^2, 3^ though our study did capture more people. Women are relatively over-represented in the FI population^27^ and may be more likely to respond to surveys, but this tendency should be equally distributed in those with and without COVID-19. Men with COVID-19 have been reported to have more severe disease than women,^28, 29^ while women may be more susceptible.^30^ Women with COVID-19 may be over-represented in our study because they had relatively milder disease, and were therefore more able to answer surveys than men. Women with PD without COVID-19 reported worsening or new onset of PD symptoms more often than men without COVID-19 in all domains except cognition. Other studies have similarly suggested that women with PD report more mood symptoms^31 32^ and fewer cognitive problems^33^ than men, and our findings may be more reflective of differences in symptoms between men and women with PD rather than reflecting changes due to the pandemic.

A recent report suggested that COVID-19 infection is more common in people homozygous for APOE ε4.^14^ Among the PwPD with genetic testing available, no one with COVID-19 had an APOE ε4 allele. Furthermore, PD-related mutations were not related to the risk of COVID-19 or symptomatic worsening during the pandemic. However, as the prevalence of these mutations is low, our sample size may not have been big enough to detect a difference in associated risk.

Among those with COVID-19, outcomes were largely similar between people with and without PD. Longer PD-duration was associated with a higher risk for poor outcomes. Similarly, a series of 10 cases identified through hospitals in Padua, Italy, and London, UK, suggested that PwPD of longer duration may be at risk of more severe illness and mortality from COVID-19.^2^ Preliminary reports from the CDC indicate that a high proportion of people with COVID-19 and neurologic disorders require hospitalization,^34^ and severity of other neurologic conditions has been associated with complications of COVID-19.^35^ Although the number of PwPD and COVID-19 studied is small, consistency among reports suggests that complications may be more frequent in advanced PwPD.

The vast majority of PwPD did not have COVID-19, yet most reported significant disruptions in many aspects of their daily lives due to the pandemic and consequent public health precautions. Longer disease duration and living alone increased the risk of disruption to medical care or other essential activities, indicating that these groups may need specific attention. Non-white race and lower household income were independently associated with difficulty obtaining medications, a particularly concerning finding that highlights the presence of barriers to healthcare access even among an online research cohort, and that these barriers are exacerbated during the pandemic. People with lower income were also less likely to find alternative means to conduct exercise and social activities and were less likely to use telemedicine. Among those with PD, lower socioeconomic status may already be associated with poor outcomes such as earlier exit from the work force^36^ and higher mortality risk.^37^ The rapid transformation of healthcare delivery increases access for many through telemedicine alternatives and will hopefully be a long-lasting consequence of the COVID-19 pandemic,^7^ but access for some already disenfranchised groups may be further reduced unless we are particularly conscious of this risk.^38^

Interruption of medical care, exercise, and social activities were also associated with exacerbation of PD symptoms. The effect was even greater among those asked to socially isolate or quarantine. In the elderly, social isolation has been associated with more rapid cognitive decline,^39^ and quarantine in particular can lead to lasting psychological effects.^40^ Individuals with PD are at high risk for anxiety,^41^ and may be particularly vulnerable to external stressors. Developing ways to minimize these adverse consequences while maintaining safety precautions are needed.

Several limitations of this study must be acknowledged. We relied on self-report of COVID-19 infection and outcomes. In sensitivity analyses, applying more stringent criteria for COVID-19 classification of cases did not change our findings. Survey completion was naturally limited to people who were healthy enough to log-in online and fill out a survey, and we likely did not capture those with very advanced PD nor those with severe COVID-19 illness. People with PD and COVID-19 may have been less likely to fill out a survey, preventing our study from identifying true differences in infection risk or outcome severity. Additionally, certain populations were underrepresented, and the fact that we did see significantly greater disruption from the pandemic in some of these groups (e.g. non-White race, lower income) indicates that the true differences may be much larger. Finally while PD-diagnosis is self-reported in FI, available data indicate high accuracy of self-reported PD in research settings; in one study that verified self-reported diagnosis of PD through use of a Webcam, a neurologist agreed with the diagnosis in 97% of cases.^42^ An effort to validate diagnoses in FI is currently underway.

Our study raises important questions for future analyses. Anosmia in COVID-19, though usually transient,^43^ may represent true viral invasion of the olfactory bulbs.^44^ Hyposmia predicts PD-associated clinical and pathological changes.^45, 46^ This association, among other neurologic manifestations in people with COVID-19,^47, 48^ has prompted worries about the possibility of SARS-CoV-2 infection triggering long-term neurodegeneration,^49^ as was observed following the 1917-1918 pandemic.^50^ Follow-up of people with and without PD who had new or worsened hyposmia may provide important insights into this question. Our survey population also provides the opportunity to investigate the long-term effects of COVID-19 on PD progression and of the social and public health aspects of the pandemic on PwPD. PwPD rely heavily on a network of outpatient healthcare, ancillary services, and social support, and interruptions of those services may have lasting implications that should be characterized longitudinally. FI has the capability to reach a broader and more diverse group than traditional, center-based research, and targeted recruitment will allow future studies to clarify the extent of and solutions for disparities in healthcare access during and after the pandemic.

In conclusion, the disruption from the COVID-19 pandemic in PD is substantial, both due to direct effects of the infection and the broader shelter-in-place guidelines. Disruptions in PD-related medical care, essential daily activities, exercise, and social activity were frequent, and contributed to worsening of motor and non-motor symptoms especially in specific at-risk groups. Compensatory activities such as telemedicine and online exercise and social programs are important, though these are not available to everyone. As the pandemic and social distancing guidelines will likely last for some time, targeted strategies should be developed to provide support to patients with all levels of resources. The COVID-19 pandemic will likely have long-term repercussions; intervention to mitigate those effects in our patients should begin as soon as possible.

## Data Availability

All survey questions and data are available for download online at: https://www.foxden.michaeljfox.org.

https://www.foxinsight.michaeljfox.org

## Acknowledgments

The authors acknowledge the many people with PD and the PD partners community who provided invaluable assistance with development and testing of the survey.

## Appendix 1

| **Name** | **Location** | **Contribution** |
| --- | --- | --- |
| Ethan G. Brown, MD | University of California San Francisco | Designed and conceptualized study; analyzed the data; drafted the manuscript for intellectual content |
| Lana M. Chahine, MD | University of Pittsburgh | Designed and conceptualized study; interpreted data; revised the manuscript for intellectual content |
| Samuel M. Goldman, MD, MPH | University of California San Francisco | Designed and conceptualized study; interpreted data; revised the manuscript for intellectual content |
| Monica Korell, MPH | University of California San Francisco | Designed and conceptualized study; major role in the acquisition of data; revised the manuscript for intellectual content |
| Emerald Mann, MPH | University of California San Francisco | Designed and conceptualized study; major role in the acquisition of data; revised the manuscript for intellectual content |
| Daniel Kinel, JD | New York, NY | Designed and conceptualized study; revised the manuscript for intellectual content |
| Vanessa Arnedo, MPH | Michael J. Fox Foundation | Designed and conceptualized study; revised the manuscript for intellectual content |
| Ken Marek, MD | Institute for Neurodegenerative Disorders | Designed and conceptualized study; revised the manuscript for intellectual content |
| Caroline M. Tanner, MD, PhD | University of California San Francisco | Design and conceptualized study; analyzed the data; drafted the manuscript for intellectual content |

## References

1. Schirinzi T, Cerroni R, Di Lazzaro G, et al. Self-reported needs of patients with Parkinson’s disease during COVID-19 emergency in Italy. Neurol Sci 2020.

2. Antonini A, Leta V, Teo J, Chaudhuri KR. Outcome of Parkinson’s Disease patients affected by COVID-19. Mov Disord 2020.

3. Cilia R, Bonvegna S, Straccia G, et al. Effects of COVID-19 on Parkinson’s disease clinical features: a community-based case-control study. Mov Disord 2020.

4. Hainque E, Grabli D. Rapid worsening in Parkinson’s disease may hide COVID-19 infection. Parkinsonism Relat Disord 2020.

5. Pantell M, Rehkopf D, Jutte D, Syme SL, Balmes J, Adler N. Social isolation: a predictor of mortality comparable to traditional clinical risk factors. Am J Public Health 2013;103:2056–2062.

6. Plagg B, Engl A, Piccoliori G, Eisendle K. Prolonged social isolation of the elderly during COVID-19: Between benefit and damage. Arch Gerontol Geriatr 2020;89:104086.

7. Bloem BR, Dorsey ER, Okun MS. The Coronavirus Disease 2019 Crisis as Catalyst for Telemedicine for Chronic Neurological Disorders. JAMA Neurol 2020.

8. Fasano A, Antonini A, Katzenschlager R, et al. Management of Advanced Therapies in Parkinson’s Disease Patients in Times of Humanitarian Crisis: The COVID-19 Experience. Mov Disord Clin Pract 2020;7:361–372.

9. Miocinovic S, Ostrem JL, Okun MS, et al. Recommendations for Deep Brain Stimulation Device Management During a Pandemic. J Parkinsons Dis 2020.

10. Garg D, Dhamija RK. The Challenge of Managing Parkinson’s Disease Patients during the COVID-19 Pandemic. Ann Indian Acad Neurol 2020;23:S24–S27.

11. Smolensky L, Amondikar N, Crawford K, et al. Fox Insight collects online, longitudinal patient-reported outcomes and genetic data on Parkinson’s disease. Sci Data 2020;7:67–67.

12. Fox Insight [online]. Available at: foxinsight.michaeljfox.org. Accessed July 8th, 2020.

13. Wallings RL, Tansey MG. LRRK2 regulation of immune-pathways and inflammatory disease. Biochem Soc Trans 2019;47:1581–1595.

14. Kuo CL, Pilling LC, Atkins JL, et al. APOE e4 genotype predicts severe COVID-19 in the UK Biobank community cohort. J Gerontol A Biol Sci Med Sci 2020.

15. D’Souza T, Rajkumar AP. Systematic review of genetic variants associated with cognitive impairment and depressive symptoms in Parkinson’s disease. Acta Neuropsychiatrica 2020;32:10–22.

16. Fox Den: A Fox Insight Research Tool [online]. Available at: foxden.michaeljfox.org. Accessed July 8th, 2020.

17. Zheng KS, Dorfman BJ, Christos PJ, et al. Clinical characteristics of exacerbations in Parkinson disease. Neurologist 2012;18:120–124.

18. Brugger F, Erro R, Balint B, Kägi G, Barone P, Bhatia KP. Why is there motor deterioration in Parkinson’s disease during systemic infections-a hypothetical view. NPJ Parkinson’s disease 2015;1:15014–15014.

19. Helms J, Kremer S, Merdji H, et al. Neurologic Features in Severe SARS-CoV-2 Infection. New England Journal of Medicine 2020;382:2268–2270.

20. Coolen T, Lolli V, Sadeghi N, et al. Early postmortem brain MRI findings in COVID-19 non-survivors. Neurology 2020.

21. Zhou L, Zhang M, Wang J, Gao J. Sars-Cov-2: Underestimated damage to nervous system. Travel Med Infect Dis 2020:101642.

22. Virhammar J, Kumlien E, Fällmar D, et al. Acute necrotizing encephalopathy with SARS-CoV-2 RNA confirmed in cerebrospinal fluid. Neurology 2020.

23. Solomon IH, Normandin E, Bhattacharyya S, et al. Neuropathological Features of Covid-19. New England Journal of Medicine 2020.

24. Chen G, Wu D, Guo W, et al. Clinical and immunological features of severe and moderate coronavirus disease 2019. The Journal of Clinical Investigation 2020;130:2620–2629.

25. Moein ST, Hashemian SM, Mansourafshar B, Khorram-Tousi A, Tabarsi P, Doty RL. Smell dysfunction: a biomarker for COVID-19. Int Forum Allergy Rhinol 2020.

26. Giacomelli A, Pezzati L, Conti F, et al. Self-reported Olfactory and Taste Disorders in Patients With Severe Acute Respiratory Coronavirus 2 Infection: A Crosssectional Study. Clinical Infectious Diseases 2020.

27. Chahine LM, Chin I, Caspell-Garcia C, et al. Comparison of an Online-Only Parkinson’s Disease Research Cohort to Cohorts Assessed In Person. J Parkinsons Dis 2020;10:677–691.

28. Qin LA-O, Li X, Shi J, et al. Gendered effects on inflammation reaction and outcome of COVID-19 patients in Wuhan. LID - 10.1002/jmv.26137 [doi].

29. Gebhard C, Regitz-Zagrosek V, Neuhauser HK, Morgan R, Klein SL. Impact of sex and gender on COVID-19 outcomes in Europe. Biol Sex Differ 2020;11:29–29.

30. Qian J, Zhao L, Ye RZ, Li XJ, Liu YL. Age-dependent gender differences of COVID-19 in mainland China: comparative study. Clin Infect Dis 2020.

31. Liu R, Umbach DM, Peddada SD, et al. Potential sex differences in nonmotor symptoms in early drug-naive Parkinson disease. Neurology 2015;84:2107.

32. Martinez-Martin P, Falup Pecurariu C, Odin P, et al. Gender-related differences in the burden of non-motor symptoms in Parkinson’s disease. Journal of Neurology 2012;259:1639–1647.

33. Reekes TH, Higginson CI, Ledbetter CR, Sathivadivel N, Zweig RM, Disbrow EA. Sex specific cognitive differences in Parkinson disease. npj Parkinson’s Disease 2020;6:7.

34. Preliminary Estimates of the Prevalence of Selected Underlying Health Conditions Among Patients with COVID-19. Morb Mortal Wkly Rep 2020;69:603 – 605.

35. Louapre C, Collongues N, Stankoff B, et al. Clinical Characteristics and Outcomes in Patients With Coronavirus Disease 2019 and Multiple Sclerosis. JAMA Neurol 2020.

36. Armstrong MJ, Gruber-Baldini AL, Reich SG, Fishman PS, Lachner C, Shulman LM. Which features of Parkinson’s disease predict earlier exit from the workforce? Parkinsonism Relat Disord 2014;20:1257–1259.

37. Yang F, Johansson AL, Pedersen NL, Fang F, Gatz M, Wirdefeldt K. Socioeconomic status in relation to Parkinson’s disease risk and mortality: A population-based prospective study. Medicine (Baltimore) 2016;95:e4337.

38. Crawford A, Serhal E. Digital Health Equity and COVID-19: The Innovation Curve Cannot Reinforce the Social Gradient of Health. J Med Internet Res 2020;22:e19361.

39. Friedler B, Crapser J, McCullough L. One is the deadliest number: the detrimental effects of social isolation on cerebrovascular diseases and cognition. Acta Neuropathol 2015;129:493–509.

40. Brooks SK, Webster RK, Smith LE, et al. The psychological impact of quarantine and how to reduce it: rapid review of the evidence. The Lancet 2020;395:912–920.

41. Broen MP, Narayen NE, Kuijf ML, Dissanayaka NN, Leentjens AF. Prevalence of anxiety in Parkinson’s disease: A systematic review and meta-analysis. Mov Disord 2016;31:1125–1133.

42. Dorsey ER, Wagner JD, Bull MT, et al. Feasibility of Virtual Research Visits in Fox Trial Finder. Journal of Parkinson’s Disease 2015;5:505–515.

43. Meini S, Suardi LR, Busoni M, Roberts AT, Fortini A. Olfactory and gustatory dysfunctions in 100 patients hospitalized for COVID-19: sex differences and recovery time in real-life. Eur Arch Otorhinolaryngol 2020.

44. Politi LS, Salsano E, Grimaldi M. Magnetic Resonance Imaging Alteration of the Brain in a Patient With Coronavirus Disease 2019 (COVID-19) and Anosmia. JAMA Neurology 2020.

45. Ross GW, Petrovitch H, Abbott RD, et al. Association of olfactory dysfunction with risk for future Parkinson’s disease. Ann Neurol 2008;63:167–173.

46. Ross GW, Abbott RD, Petrovitch H, et al. Association of olfactory dysfunction with incidental Lewy bodies. Mov Disord 2006;21:2062–2067.

47. Pleasure SJ, Green AJ, Josephson SA. The Spectrum of Neurologic Disease in the Severe Acute Respiratory Syndrome Coronavirus 2 Pandemic Infection: Neurologists Move to the Frontlines. JAMA Neurol 2020.

48. Romero-Sánchez CM, Díaz-Maroto I, Fernández-Díaz E, et al. Neurologic manifestations in hospitalized patients with COVID-19: The ALBACOVID registry. Neurology 2020.

49. De Felice FG, Tovar-Moll F, Moll J, Munoz DP, Ferreira ST. Severe Acute Respiratory Syndrome Coronavirus 2 (SARS-CoV-2) and the Central Nervous System. Trends in Neurosciences 2020.

50. Hoffman LA, Vilensky JA. Encephalitis lethargica: 100 years after the epidemic. Brain 2017;140:2246–2251.

